# Safety and Procedural Success of Minor Salivary Gland Biopsy in Sjögren’s Disease: A Retrospective Analysis of 202 Patients

**DOI:** 10.1101/2025.06.25.25330262

**Authors:** Georg A. Ehart, Daniel Pietsch, Sabine Zenz, Josef Hermann, Christian Dejaco, Jens Thiel, Martin H. Stradner

## Abstract

**Objective:** Minor salivary gland biopsy (MSGB) is a standard procedure in the diagnosis of Sjögren’s disease (SjD). Biopsy techniques using a vertical or horizontal mucosal incision are applied in daily practice; the former may be safer due to underlying neuroanatomy. Here we aim to quantify and classify adverse events (AEs) of MSGB using vertical incision technique.

**Patients and Methods:** Medical records of patients with suspected SjD who underwent MSGB at our outpatient clinic between January 2014 and April 2024 and had follow-up visits were analyzed. The MSBG consisted of a 1 cm linear vertical incision in the oral mucosa of the lower lip to collect up to five lip salivary glands.

**Results:** 202 of 257 patients undergoing MSGB met inclusion criteria. Median follow-up time was 16.5 (0.5-124.0) months. Four biopsies (2%) did not yield sufficient material for histological analysis. One serious adverse event (0.5%), local persistent paresthesia, and two moderate adverse events (1.0%), including one case of prolonged bleeding and one unclassified event, were reported. Mild AEs occurred in 18 patients (8.9%). Local discomfort after biopsy affected 32 patients (15.8%). AEs subsided either during or up to 2 hours after biopsy in 14 patients (6.9%) and in 38 patients (18.8%) within three weeks, respectively.

**Conclusions:** MSGB with vertical incision technique has a very low frequency of serious and long-term complications with a success rate of 98%.

**Significance and Innovations:**

- Minor salivary gland biopsy with lateral vertical incision technique has a success rate of 98%.
- Serious complications occurred less frequent than previously reported for horizontal incision technique.

## Introduction

Sjögren’s disease (SjD) is a systemic autoimmune disease primarily characterized by involvement of exocrine glands typically presenting with dryness of the mouth (xerostomia) and eyes (xerophthalmia) as well as extra-glandular manifestations [1]. To classify SjD according to the current ACR/EULAR classification criteria either detection of Anti-SSA/Ro antibodies or histopathological examination of salivary glands with focal lymphocytic sialadenitis and a focus score of ≥ 1 foci/4 mm^2^ is required [2]. In order to obtain salivary gland tissue MSGB, first described by Chisholm and Mason in 1968, is a widely used method for assessing patients with suspected SjD [3]. Further indications for MSGB include the diagnostic workup of sarcoidosis and amyloidosis [4]. Several techniques have been proposed with variations in incision-shape (elliptical, longitudinal), - length (2-30 mm) and - orientation (vertical, horizontal, oblique) [3, 5-22]. A punch biopsy technique has also been described [23, 24]. While the procedure is widely practiced, it is not without risk. Among several possible complications nerve lesions are considered to be the most relevant due to the risk of a permanent neurological deficit [25]. Some studies report up to 6% of permanent paresthesia following MSGB [10]. Compared to conventional horizontal incision, linear vertical incision may be a safer biopsy technique as the incision is more parallel to the branches of the mental nerve [26]. However, only three smaller studies have investigated the risk and usefulness of a vertical incision in order to obtain labial salivary gland tissue [9, 22], one of which only examines its application in the diagnosis of neonatal hemochromatosis [17]. Furthermore exact assessment of adverse events has been prevented by lack of standardized definition and categorization of surgical complications [27]. Here we aimed to quantify and classify adverse events (AEs) of MSGB using vertical incisions in a large retrospectively analyzed cohort.

## Methods

### Patients

We conducted a retrospective analysis of all patients undergoing MSGB between January 2014 and April 2024 at the Division of Rheumatology and Immunology, Medical University of Graz. Inclusion criteria were 1) age ≥ 18 years at the time of biopsy, 2) presence of biopsy documentation, 3) availability of histopathological examination records, 4) at least one clinic visit during follow-up with documentation of AE assessment, and 5) biopsy performed because of suspected SjD. No exclusion criteria were applied.

### Data collection

Ethical approval for this retrospective study was obtained from the Institutional Review Board of the Medical University of Graz, in accordance with the Declaration of Helsinki. All data were extracted from the database of the Division of Rheumatology and Immunology. Demographic characteristics included gender and age at time of biopsy. In addition, medical history and current medication at time of biopsy as well as clinical parameters were collected as follows: xerostomia and xerophthalmia, Raynaud’s phenomenon and parotid swelling (past or present), results of unstimulated whole saliva flow, Saxon Test, Schirmer test, ESSDAI, ESSPRI, OSDI, Xerostomia Inventory and FACIT. Finally, time of follow-up, diagnosis of SjD, diagnosis of other autoimmune diseases, focus score and highest ESSDAI during follow-up were extracted. Patients were followed from the time of biopsy until the most recent documented contact. All included patients underwent a minimum two-week follow-up, in accordance with a schedule requiring an outpatient visit at least two weeks post-procedure to discuss biopsy findings and record adverse events as necessary. Follow-up usually ended at that point if patients were found not to have a rheumatic disease.

### Biopsy technique

All patients had provided written informed consent for the biopsy before the procedure. Biopsies were performed by one of five experienced rheumatologists or immunologists (JH, MS, CD, SZ, and DP) with the patient sitting in an upright position. After application of anesthesia (2% lidocaine, 0.0005% epinephrine) a superficial vertical incision of 1 cm in length, incising only the mucosa, was made 2-3cm lateral to the midline of the lip. The glands were dissected carefully under direct vision. Up to five glands were then collected using scalpel and tweezer. The wound was left to heal spontaneously without any sutures. After the procedure patients were asked to stay in the clinic for two hours during which they were instructed to perform mild compression of the lip for about 30 minutes. Patients were routinely asked to report sequelae after biopsy at the follow-up visit.

### Classification of complications

Based on the NIA guidelines for AEs and serious AEs [28] we classified complications regarding severity into (1) serious AE (death, life-threatening situations, disability or permanent damage, AE that requires hospitalization or intervention to prevent permanent damage), (2) moderate AE (events that lead to termination of biopsy – e.g. bleeding or vasovagal reaction – disability or damage persisting for longer than 3 weeks with full restitution over the course), (3) mild AE (symptoms of minor irritation which do not require medical intervention; signs and symptoms are transient persisting no longer than two weeks) and (4) local discomfort (up to two weeks after biopsy). As local discomfort is to be expected after the procedure, AEs classified as such were not considered complications.

Furthermore, AEs were classified into (1) immediate AE (during biopsy or within two hours after the procedure), (2) medium-term AE (persisting up to three weeks post-procedure) and (3) long-term AE (persisting for longer than three weeks post-procedure). Immediate AEs were recorded during the two-hour post-procedure period in the clinic, medium-term AEs at the first outpatient visit, and long-term AEs during subsequent follow-up in patients diagnosed with SjD.

Causality was classified in (1) definitely related AE (the AE is clearly related to the biopsy; AE that follows a reasonable temporal sequence from administration of the study intervention, follows a known or expected response pattern to the biopsy and that could not be reasonably explained by the known characteristics of the subject’s clinical state), (2) possibly related AE (AE that follows a reasonable temporal sequence from biopsy follows a known or expected response pattern to the suspected intervention, but that could readily have been produced by a number of other factors) and (3) not related AE (AE, that is clearly not related to the biopsy; another cause of the event is most plausible; clinically plausible temporal sequence is inconsistent with the onset of the event and the biopsy; Causal relationship is considered biologically implausible).

### Data processing and statistical analysis

Descriptive statistics were used to summarize and subsequently analyze the data. A significance level of 5% was set. Parameters were tested for normal distribution using the Shapiro-Wilk test. Normally distributed data are presented as mean ± standard deviation (mean ± SD). Non-normally distributed data are described using the median, minimum and maximum (median; minimum–maximum) and were compared using the Mann-Whitney-U-test and the Kruskal-Wallis-test. The t-test was used to compare normally distributed data; categorical data were compared using the chi-square test. The statistical analysis of the data was performed with IBM® SPSS® Statistics Version 29.0.1.0 (Armonk, New York: IBM Corp).

## Results

### Patients

Among the 257 patients who underwent MSGB, 202 met the inclusion criteria. The majority of patients were women (86.1%). Mean age was 54.6 years (± 14.2). 15.5% had an established systemic autoimmune disease at time of biopsy. 79.3% reported xerostomia, 83.8% xerophthalmia, 13.6% had Raynaud’s phenomenon and 12.1% reported parotid swelling in the past or present. Mean ESSPRI was 5.1 (± 2.5) and OSDI 43.8 (± 24.3). Median ESSDAI was 2 (0-24), Schirmer test result was 3 (0-37.5) mm, Xerostomia Inventory 2.1 (0.5-4), FACIT 2.2 (0-7) and Focus-Score 0.46 (0–9). Median follow-up time was 16.5 months (0.5-124) during which 59.9% received the diagnosis of SjD.

### Biopsy results

44.4% had a focus score ≥ 1 according to Fisher et al. [29]. In four biopsies (2%) sparse or no salivary gland tissue in the specimen was detectable, thus preventing calculation of the focus score. In two cases the biopsy had to be terminated due to an AE, though enough specimen for valid assessment could be collected.

### Adverse Events

In 43 cases (21.3%) one AE and in 5 cases (2.5%) two events were reported. Categorized according to severity most AEs (62.3%) were classified as local discomfort, with a median duration of 3 days (1-14). Regarding local discomfort as an expected AE, this means that 9.4% of all patients suffered from a complication. The full data is presented in Table 1. The most common events were bleeding and vasovagal reaction, both accounting for 11.3% of all adverse events. Vasovagal reactions were all immediate AEs, among the six cases of bleeding, 5 were immediate and one was medium-term. Table 2 summarizes recorded AEs. The three events classified as “others” were perforation of the lower lip during application of anesthesia, one case of hyperventilation and one case of perioral redness and sensation of heat, which quickly resolved after termination of the procedure. All AEs where definetely related to the biopsy.

**Table 1.**
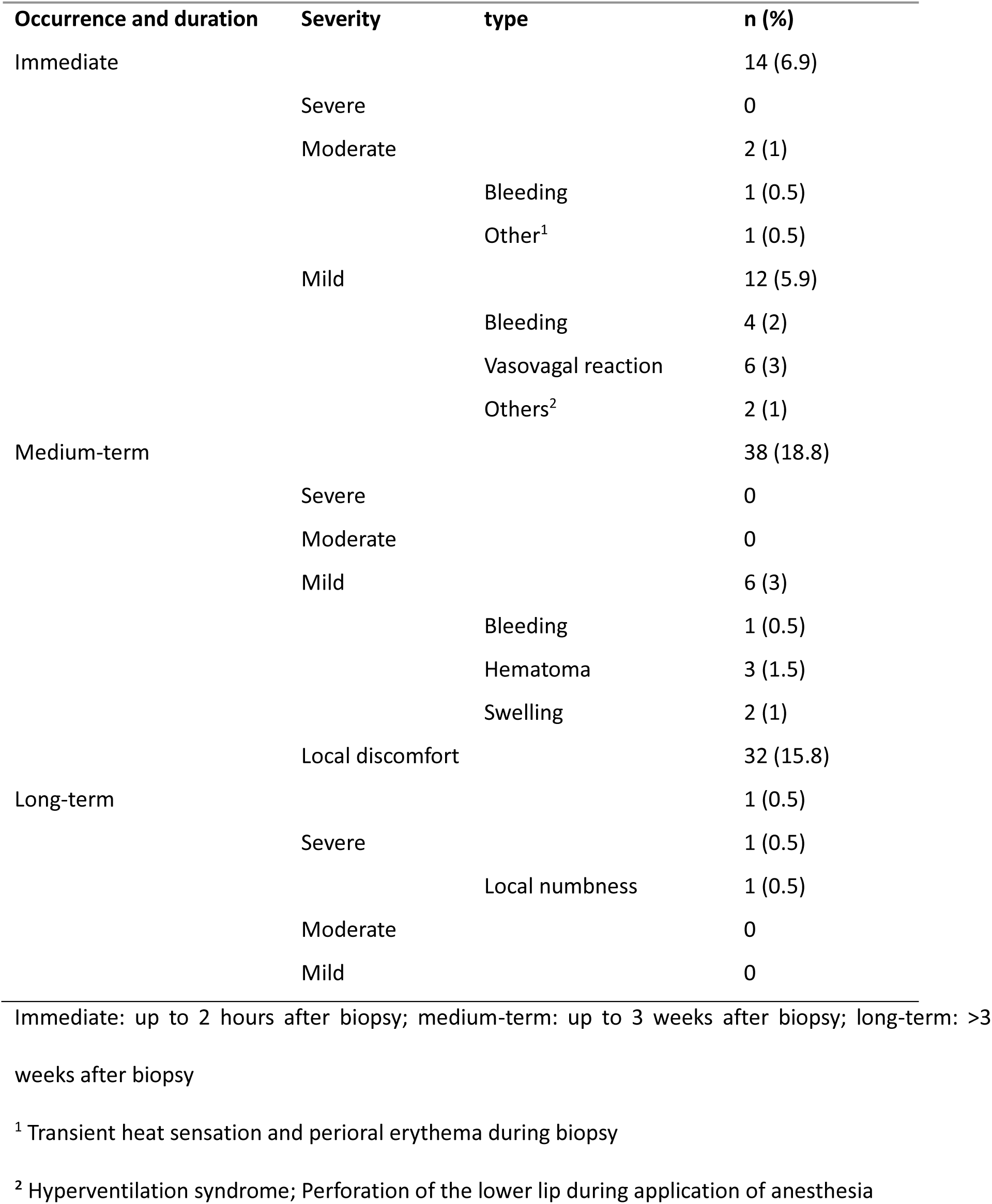
Adverse events reported.

**Table 2.**
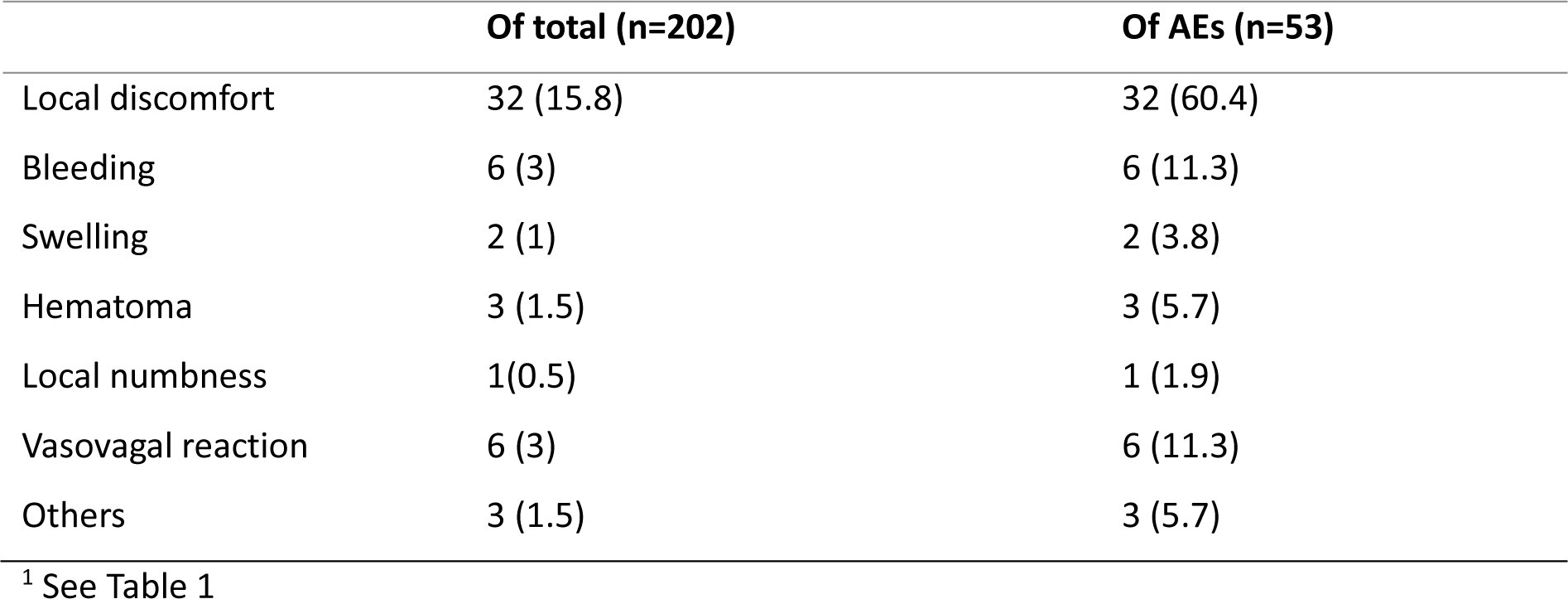
Types of events.

### Risk factors

Occurrence of AEs was associated with subjective xerophthalmia (p=0.036), although this could not be veryfied by comparing mean Schirmer test results between patients with and without a reported AE (p=0.497). Diagnosis of SjD was not associated with the occurence of AEs (p=0.933), neither was the diagnosis of other sytemic autoimmune diseses (p= 0.896). The data are presented in Table 3.

**Table 3.**
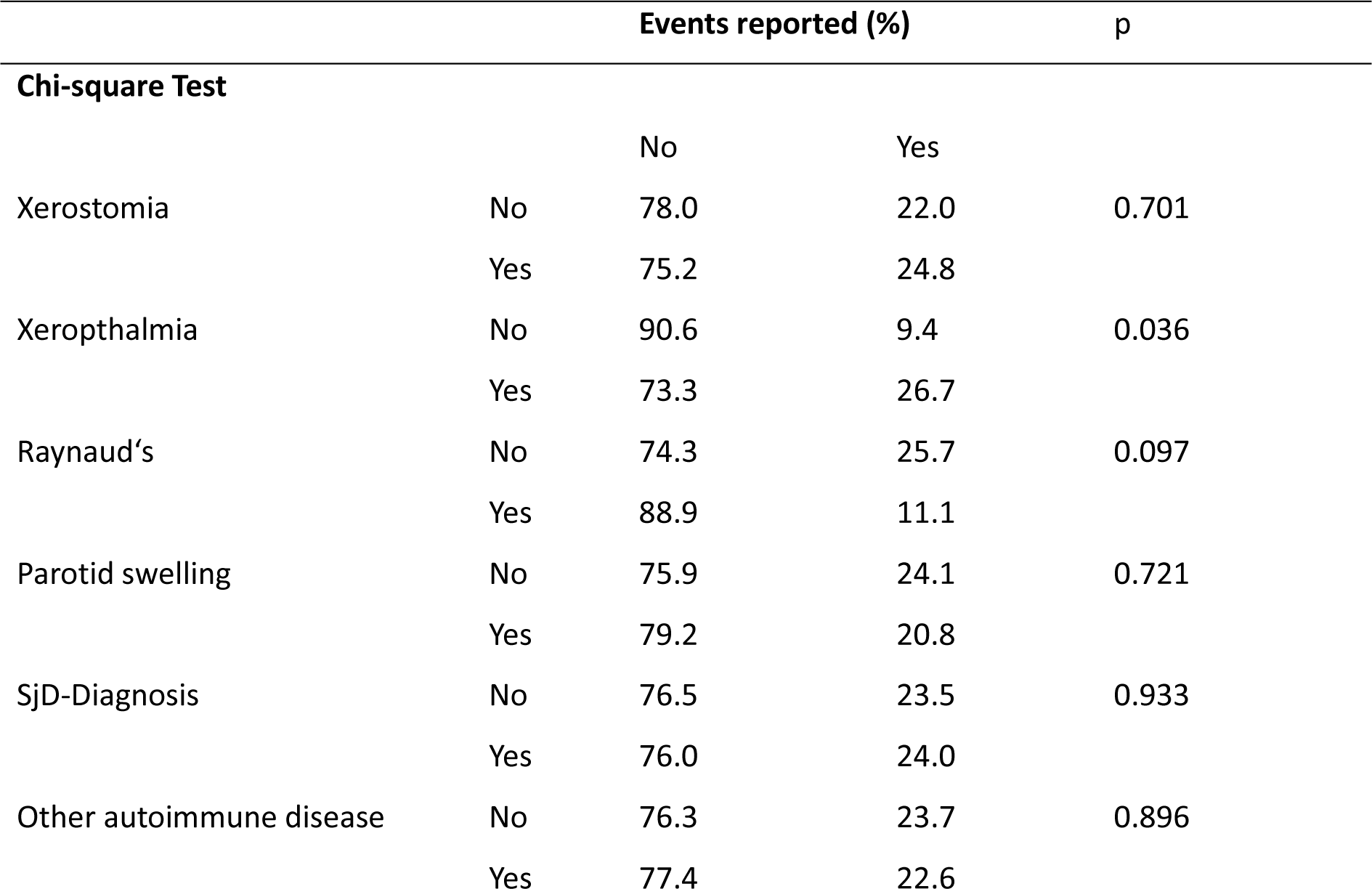
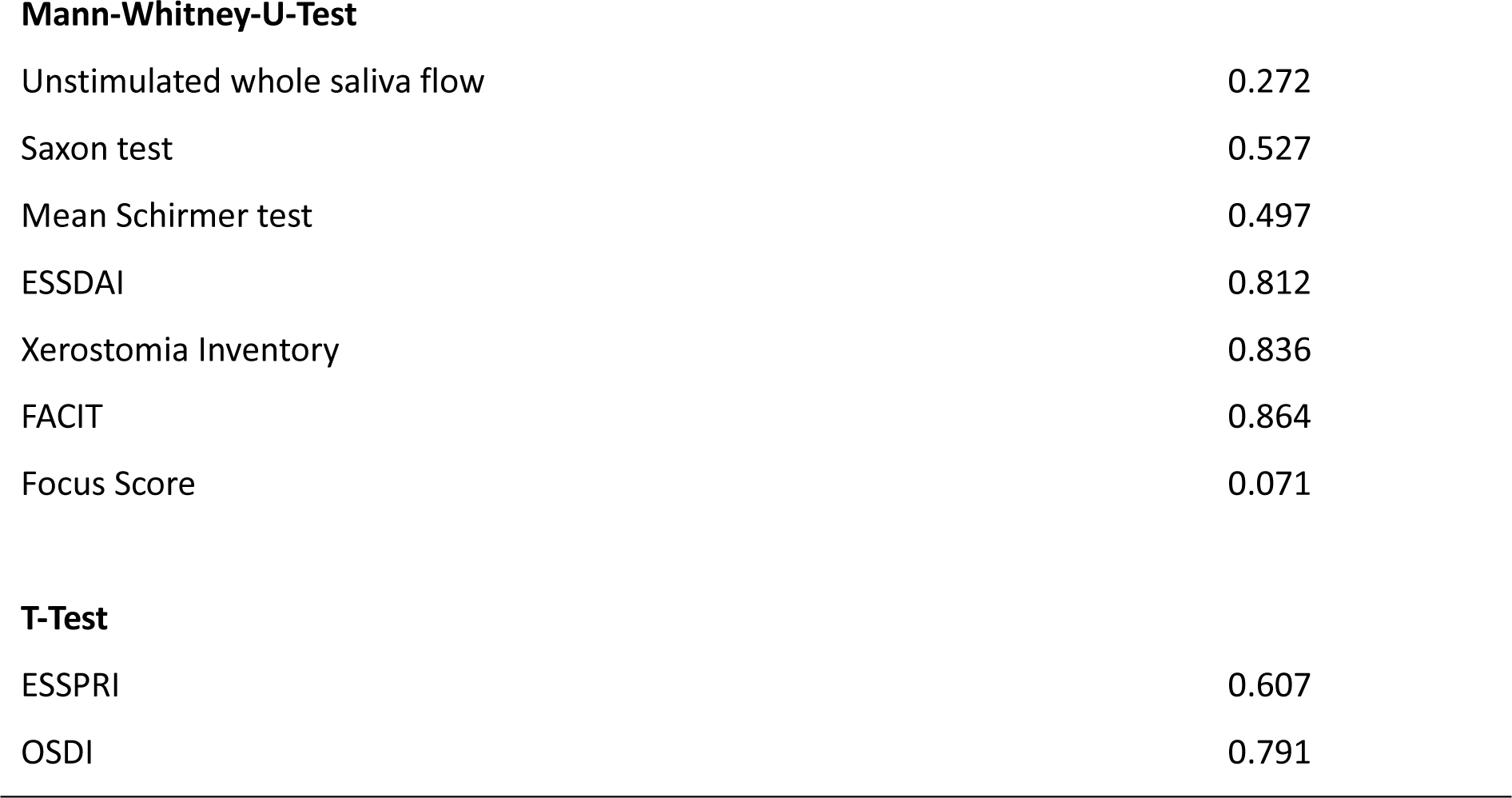
Statistical Tests.

|  |  | Events reported (%) |  | p |
| --- | --- | --- | --- | --- |
| Chi-square Test |  |  |  |  |
|  |  | No | Yes |  |
| Xerostomia | No | 78.0 | 22.0 | 0.701 |
|  | Yes | 75.2 | 24.8 |  |
| Xerophthalmia | No | 90.6 | 9.4 | 0.036 |
|  | Yes | 73.3 | 26.7 |  |
| Raynaud’s | No | 74.3 | 25.7 | 0.097 |
|  | Yes | 88.9 | 11.1 |  |
| Parotid swelling | No | 75.9 | 24.1 | 0.721 |
|  | Yes | 79.2 | 20.8 |  |
| SjD-Diagnosis | No | 76.5 | 23.5 | 0.933 |
|  | Yes | 76.0 | 24.0 |  |
| Other autoimmune disease | No | 76.3 | 23.7 | 0.896 |
|  | Yes | 77.4 | 22.6 |  |

**Mann-Whitney-U-Test**
|  |  |
| --- | --- |
| Unstimulated whole saliva flow | 0.272 |
| Saxon test | 0.527 |
| Mean Schirmer test | 0.497 |
| ESSDAI | 0.812 |
| Xerostomia Inventory | 0.836 |
| FACIT | 0.864 |
| Focus Score | 0.071 |

**T-Test**
|  |  |
| --- | --- |
| ESSPRI | 0.607 |
| OSDI | 0.791 |

## Discussion

Here we report the largest cohort of patients undergoing MSGB with mucosal vertical incision technique. The MSGB technique was safe (one severe AE in 202 cases) and yielded sufficient specimen for histological analysis in 98% of the cases. This may be an advantage especially compared to punch biopsy techniques, where tissue is harvested blindly, although the two known studies on punch biopsies do not state the success rate of their sampling [23, 24]. Another advantage seems to be a very low frequency of labial sensory defects. Only one patient (0.5%) reported ongoing local numbness after the procedure. This incidence is three times lower than the pooled risk for neurological AEs in biopsies with linear incisions estimated by Varela Centelles et al. (1.45%) [25] and six times lower than the one reported by Valdez et al. (3%) [30]. We believe this difference is due to the fact that most authors with a linear incision technique use a non-vertical incision technique. [5-7, 10, 13, 16, 18-20]. In contrast, the only study directly comparing a vertical versus a horizontal incision technique reported no significant difference between these two approaches regarding paresthesia. However, a vertical incision was correlated with a lower incidence of pain, swelling, scar formation and difficulty in eating when compared to a horizontal incision technique. Finally, the only two cases of persisting paresthesia were reported in the horizontal incision group [22]. The cutaneous branches of the mental nerve run vertically to slightly oblique ventral of the lip salivary glands in most cases [26] (Figure 1 A and B). This could explain the lower risk of nerve injury using vertical incision technique. In approximately 10% branches of the mental nerve run horizontally [26] (Figure 1 C). Injury to this variant may have been the cause of persistent local paresthesia of the lip in our patient. When preventing neuronal injury, depth of the incision may even be more important than its orientation. A superficial incision cutting only the mucosa was sufficient to access the salivary glands, but does not reach the branches of the mental nerve running ventral of the glands [26].

**Figure 1.**
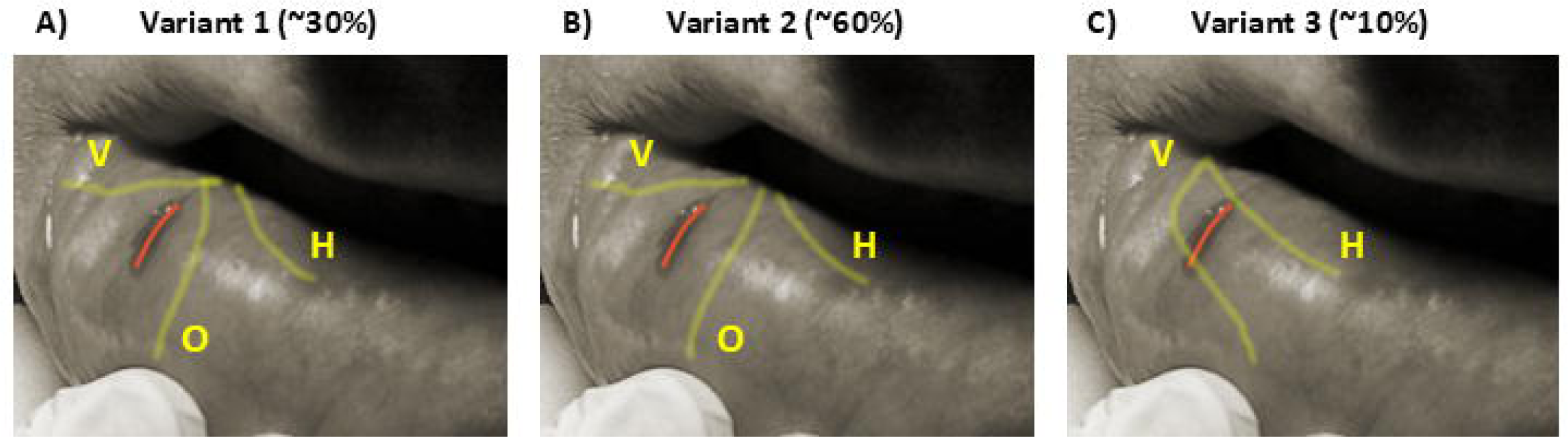
Vertical incision and location of the branches of the mental nerve. Photograph of a minor salivary gland biopsy overlayed with diagrams of mental nerve branches according to Alsaad et al. [26]. Red line, biopsy incision. Yellow lines, branches of the mental nerve. V, vertical branch; O, oblique branch; H, horizontal branch.

The overall complication rate observed in this study was 9.4%, which is similar to the combined prevalence of postsurgical complications estimated by Valdez et al. (11%) [30]. Local discomfort after the biopsy was the most common AE, with 15.8% of all patients affected. This incidence is significantly higher than the given incidence of pain in a study describing a minimally invasive technique by Caporali et al. (7.32%) [11, 21]. Another study describing a technique of 5-7mm incisions described local mediate term pain occurring in 3 of 118 patients. [8] A possible explanation for this difference could be the larger incision required by the technique described in this study.

The retrospective, single-center design of this study imposes inherent limitations. Selection bias may have occurred because patients with a documented AE are more likely to be included, potentially skewing the results. The absence of a prospective data collection process means some AEs could have been underreported or missed, especially if not documented systematically. Additionally, the single-center setting limits the generalizability of the findings, as practice patterns and patient populations may differ elsewhere. The lack of a prospective comparator arm - such as patients undergoing alternative MSGB techniques (e.g., horizontal incision or punch) - restricts the ability to directly compare outcomes and safety profiles across different methods. These limitations highlight the need for future multicenter, prospective, randomized studies to validate the findings and provide more comprehensive evidence.

Furthermore, ascertainment and recall bias for medium-term AEs primarily arises from reliance on patient self-reporting and clinical assessment during follow-up. Because this depends on patients’ memory and judgment of symptom relevance, some medium-term AEs may be underreported, potentially biasing AE incidence estimates. To mitigate this, only patients attending at least one outpatient visit - generally two weeks post-procedure - were included. These visits allowed clinicians to actively inquire about AEs, reducing reliance on unprompted patient recall. Immediate AEs were directly observed in the clinic within two hours post-procedure, minimizing underreporting for this period. Nonetheless, especially some medium-term AEs may still have gone unreported.

As 40.1% of patients were ultimately not diagnosed with a rheumatic disease, follow-up in this cohort typically concluded after the outpatient visit to discuss biopsy results, which generally occurred two weeks post-procedure. Consequently, the incidence of long-term AEs occurring beyond this timepoint could not be determined. Given that local numbness - the only anticipated long-term AE following MSGB - would be expected to have manifested by this time, we consider the number of missed AEs to be negligible.

## Conclusion

We conclude that the MSGB technique described in this study is a safe and useful approach with a low incidence of neurological complications and a high success rate. Considering the threefold lower incidence of neurological complications compared to biopsy techniqes with horizontal linear incisions we recommend a linear vertical incision of the mucosa. More prospective studies directly comparing different techniques should be carried out to confirm these results and to further investigate long-term complications.

## Funding (if specific to this study)

none

## Conflict of interest statement (in addition to author disclosure form)

none

## Data availability statement

Full data available from the authors upon request.

## Notes

### Competing Interest Statement

The authors have declared no competing interest.

### Funding Statement

This study did not receive any funding

### Author Declarations

Ethics committee of Mical University of Graz gave ethical approval for this work

### Summary of Updates

A correction in the title was made

